# Effect of Baduanjin exercise of obstructive sleep apnea-hypopnea in chronic obstructive pulmonary disease: a study protocol for a randomized, single-blind, controlled trial

**DOI:** 10.1101/2023.09.21.23295889

**Authors:** Xi Zhang, Yi-hang Cai, Kang-Jie Ye, Yu-Jin Li, Ji-Qiang Li, Zhong-De Zhang

## Abstract

**Background:** Baduanjin is one of the traditional Chinese treatments for respiratory diseases. However, its effects on overlap syndrome (obstructive sleep apnea-hypopnea and chronic obstructive pulmonary disease) have not been rigorously tested. This study aims to evaluate the benefits and safety of Baduanjin treatment in chronic obstructive pulmonary disease (COPD) patients with obstructive sleep apnea-hypopnea (OSAH).

**Methods:** COPD patients with OSAH will be randomly allocated in a 1:1 ratio to Baduanjin group or control group. Both groups receive continuous positive airway pressure therapy and basic medication for 12 weeks. Baduanjin group will receive additional instructional Baduanjin exercises. The primary outcomes are BODE index. The secondary outcomes are polysomnogram, cardiopulmonary exercise test, Beck Depression Scale-II, and Saint George’s Respiratory Questionnaire.

**Discussion:** This trial will provide preliminary evidence about the efficacy and safety of Baduanjin exercise for COPD patients with OSAH. Baduanjin exercises may become an additional option for pulmonary rehabilitation of OSA-COPD overlap syndrome.

**Trial registration:** ClinicalTrials.gov, ID: NCT ChiCTR2200063171. Registered retrospectively on 1 June 2022.

## 1. Background

OSA-COPD overlap syndrome describes the combination of obstructive sleep apnea-hypopnea (OSAH) and chronic obstructive pulmonary disease (COPD) ^1^. The prevalence of COPD is approximately 10% among adults ^2^, and OSAH has a prevalence ranging from 9 to 38%^3^. Therefore, it is not uncommon to have OSA-COPD overlap syndrome. OSAH and COPD are comorbid in a range from 20% to 55%^4-6^, and patients with moderate to severe COPD may have a higher prevalence^7^. Both COPD and OSAH are linked to a number of physiological and biological disturbances, including hypoxia and inflammation, which may contribute to cardiac disease and other comorbidities^8^. OSAH may exacerbate hypoxemia due to COPD during sleep, further worsening prognosis and symptoms. People with symptoms of OSAH (snoring, witnessed apnoea, unrefreshing sleep, waking headaches, excessive sleepiness, impairment), diagnosis of COPD and features of hypoventilation such as waking headaches, peripheral edema, hypoxemia (low oxygen saturation < 94% on air) and unexplained polycythaemia have a high probability of OSA-COPD overlap syndrome and should be assessed appropriately ^9^.Treatment for this syndrome consists mainly of dietary measures, weight loss, and exercise, and continuous positive airway pressure (CPAP).Treatment efficacies are affected by patient compliance with the treatment and the severity of the disease.

Baduanjin, or Eight-section Exercises, is an important component of TCM pulmonary rehabilitation. As a form of traditional Chinese health and fitness Qigong exercise, Baduanjin has a long history dating back to the Song Dynasty (960-1279). With easy postures, Baduanjin emphasizes the coordination of movement, breathing and intention. This Qigong is widely used in chronic obstructive pulmonary disease, diabetes, chronic heart disease, insomnia and other diseases, with the effect of improving cardiopulmonary endurance, relieving tension and anxiety, improving sleep quality^10-15^. Besides, Baduanjin is also recommended as a therapy for the recovery phase of novel coronavirus pneumonia.^16,17^. However, the effectiveness of Baduanjin exercise for OSA-COPD overlap syndrome lacks high-quality evidence. Therefore, we conduct this randomized, single-blind, controlled trial to evaluate the benefits and safety of Baduanjin exercise in COPD patients with OSAH.

## 2. Methods/Design

The study is a randomized, single-blind (assessor), parallel trial conducted at the Guangdong Provincial Hospital of Chinese Medicine. Participants will be recruited from September, 2022 through December, 2023. Participants will be randomly allocated in a 1:1 ratio to Baduanjin group or control group. The study period is 3 months. In addition to continuous positive airway pressure therapy and conventional therapy, patients in Baduanjin group will participate in a 30-min Baduanjin exercise session four times a week for 12 weeks. The study is approved by our institutional research ethics boards (ID BF2022-139-1), and all patients provide written informed consent. If there is any amendment to the protocol, approval will be sought from the Ethics Committee. The trial was registered at ClinicalTrials.gov (NCT ChiCTR2200063171), and the trial will be performed in accordance with the principles of the Declaration of Helsinki and Good Clinical Practice guidelines. Study flow of the trial is depicted in **Figure 1**. The schedule of trial enrollment, interventions and assessments is provided in **Table 1**.

**Figure 1.**
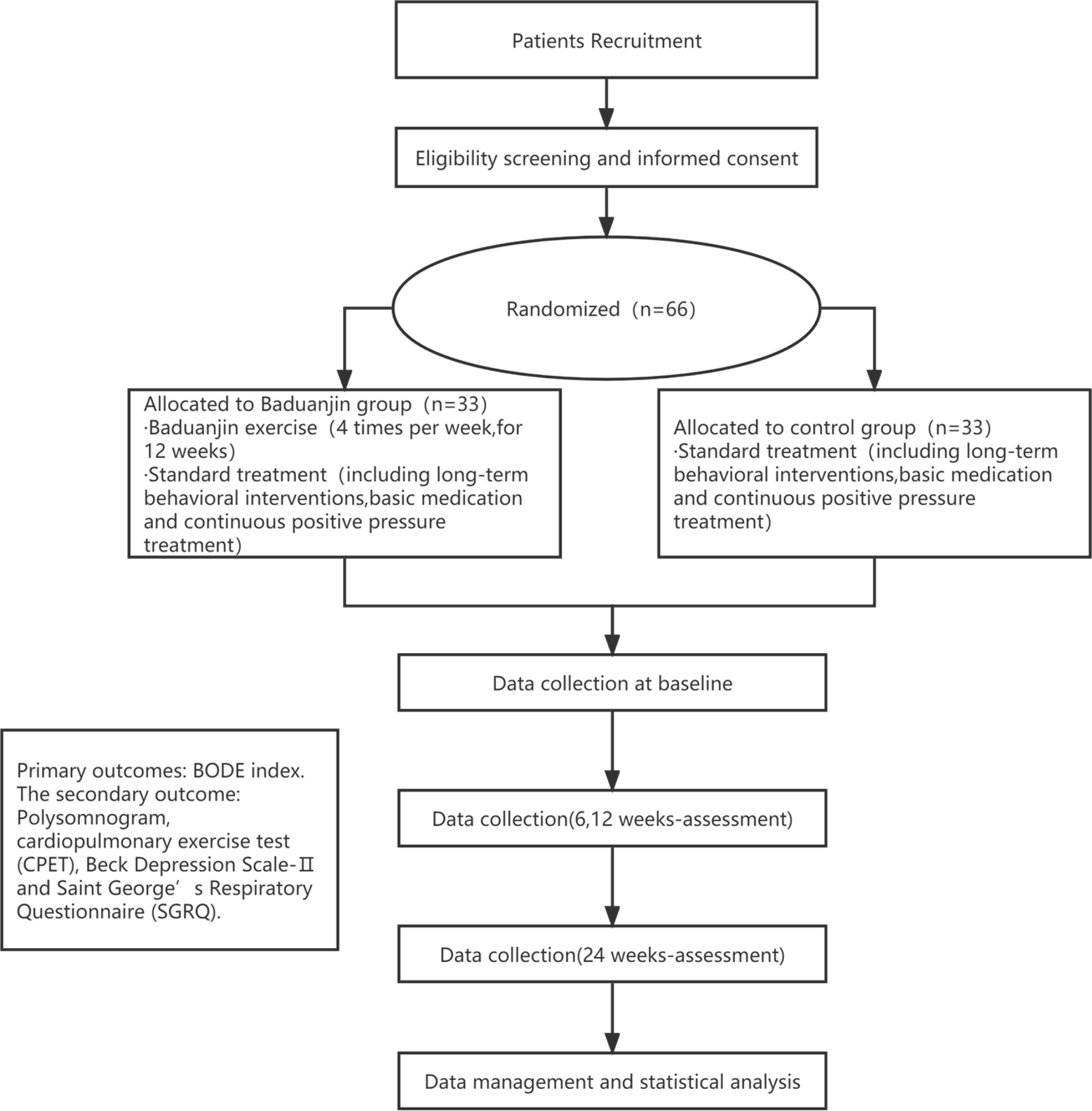
Schedule of trial enrollment, interventions and assessments

**Table 1.**
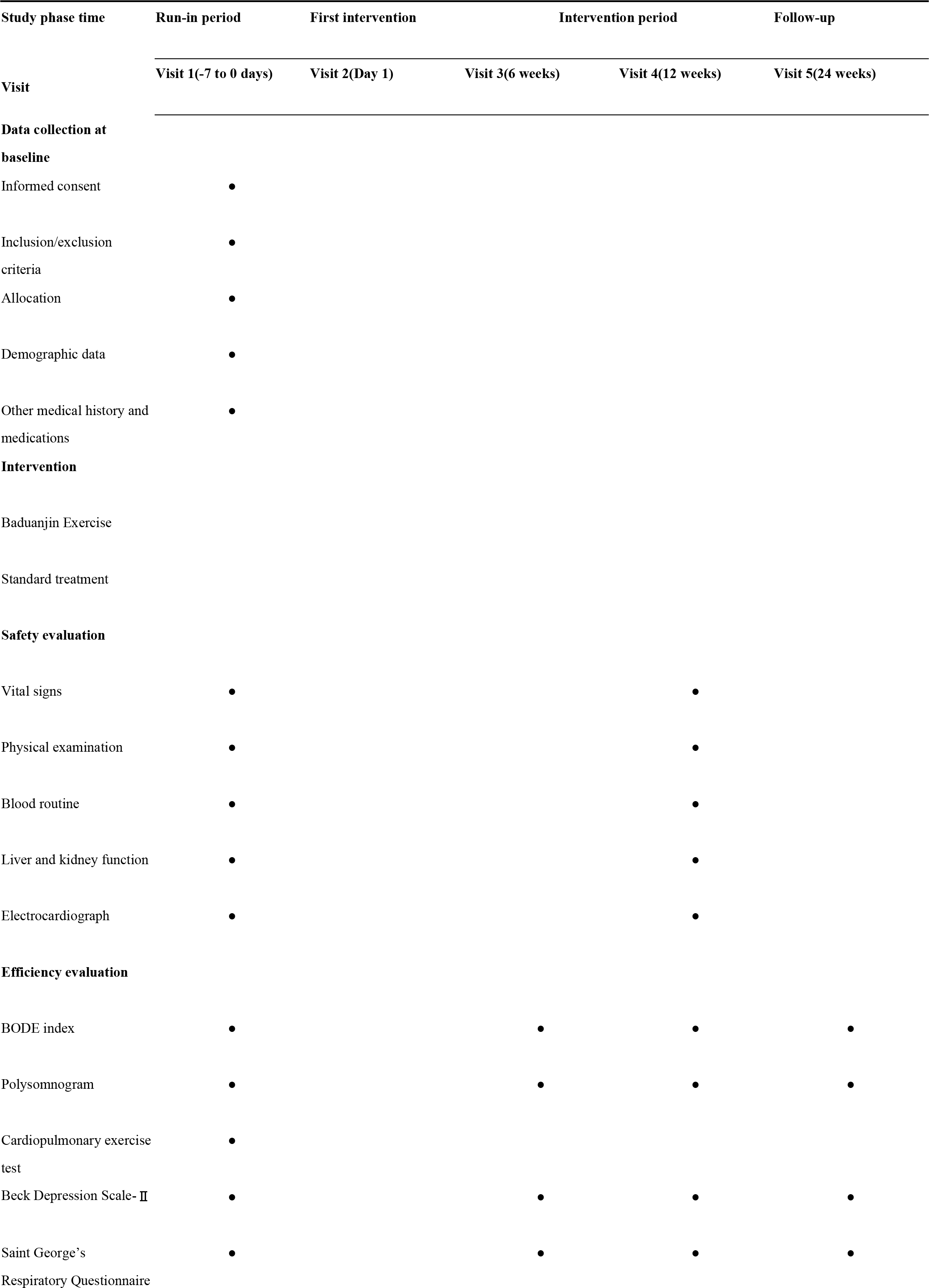
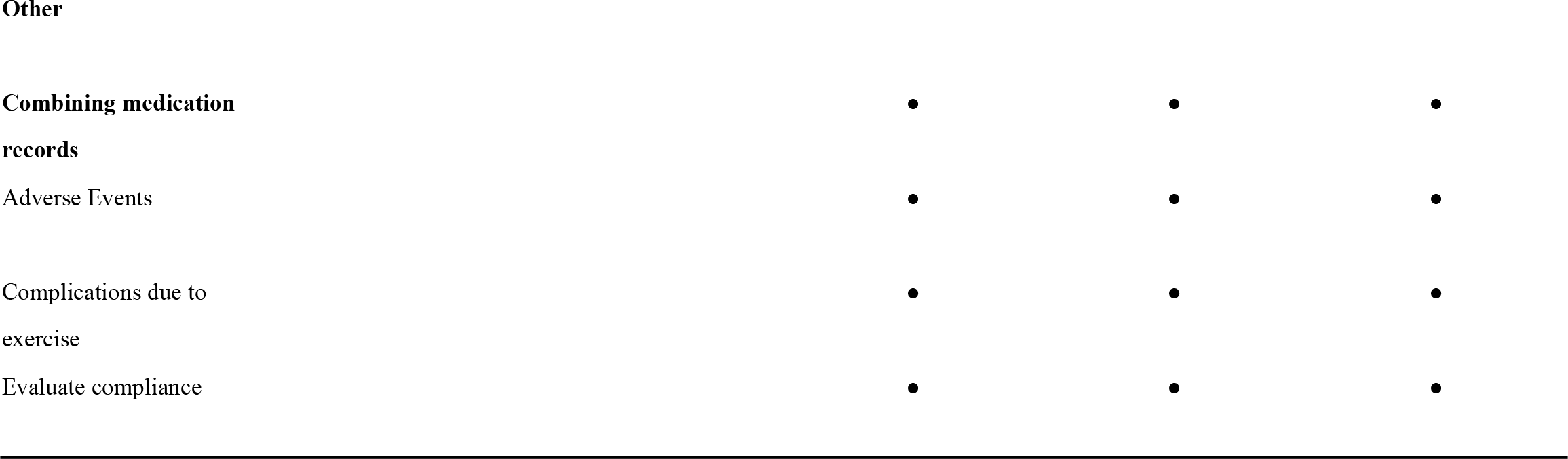
Study schedule.

### 2.1 Inclusion criteria

1. Stable COPD diagnosed by *Guidelines for Diagnosis and Management of Chronic Obstructive Pulmonary Disease* (revised version 2021)^18^, with severity class II-IV.
2. OSAH diagnosed by *Clinical Practice Guideline for Diagnostic Testing for Adult Obstructive Sleep Apnea: An American Academy of Sleep Medicine Clinical Practice Guideline*^19^, with apnea hypopnea index ≥30 events/hour.
3. Age ≥40 years, males or females.
4. Written informed consent before randomization.

### 2.2 Exclusion criteria

1. recent COPD episode (within 1 month).
2. unable to perform rehabilitation exercises.
3. participated in Baduanjin or other similar types of complementary and alternative therapy within the past 3 months.
4. poor compliance may not be adherent to the study judged by the investigator (due to the difficulty of management or other reasons).

### 2.3 Interventions

#### Control group

Patients in the control group are informed to maintain a standard treatment for 3 months according to the following protocol.

1. Long-term behavioral interventions: weight control, smoking cessation, alcohol cessation, avoidance of sedative-hypnotics before sleep, postural therapy.
2. Basic medication: standardized use of antibacterial drugs, bronchodilators, expectorants and glucocorticoids according to individual differences.
3. Continuous positive pressure treatment: continuous positive pressure ventilation via nose or face mask. Routine IPAP is set at 15-20 mmHg, EPAP is set at 8-12 mmHg, and respiratory therapy is routinely worn at night and adjusted according to the patient’s polysomnogram report and blood gas analysis.

#### Baduanjin group

In addition to the standard treatment above, a 12-week Baduanjin intervention is provided to patients in the Baduanjin group. In the 2 weeks prior to the intervention, professional instructors from Guangzhou University of Chinese Medicine will teach patients Baduanjin movements to ensure that the intervention is standardized. Patients will also receive videos of Fitness Qigong Baduanjin, compiled by the Health Qigong Management Center of the National Sports. Baduanjin consists of the following eight movements: (1) Holding the hands high with palms up to regulate the internal organs; (2) Posing as an archer shooting both left- and right-handed; (3) Holding one arm aloft to regulate the functions of the spleen and stomach; (4) Looking backwards to prevent sickness and strain; (5) Swinging the head and lowering the body to relieve stress; (6) Moving the hands down the back and legs, and touching the feet to strengthen the kidneys; (7) Thrusting the fists and making the eyes glare to enhance strength; (8) Raising and lowering the heels to cure diseases.

The Baduanjin program requires 30-minute exercise sessions, four times a week for 12 weeks, based on the patient’s physical fitness. During the 12-week intervention period, researchers will track and record participation of patients to determine whether they have attended all of the scheduled sessions. Patients who miss a class will be required to attend a makeup class. Documentation of the compliance percentage will be included in the CRF. The rate of Baduanjin compliance = (total number of times planned − number of absence) / total number of times × 100%. Compliance rates of greater than 80% are considered as good. Compliance rates of less than 80% are considered as poor. Patients will be asked to participate in follow-up visits regularly according to the study protocol, and researchers will inform them of the follow-up schedule in advance. Compliance rates of less than 20% is considered as dropout.

### 2.4 Study outcome measures

The primary and secondary outcomes will be assessed at baseline (−2 to −1 week), intermediate stage (6 weeks), the end of the intervention (12 weeks) and the end of the additional 12-week follow-up period (24 weeks). All measurements are conducted by the professional operators who do not know the allocation group at the Guangdong Provincial Hospital of Chinese Medicine.

The primary outcome measures are BODE index. The BODE index is a multidimensional index of disease severity in COPD, incorporating four independent predictors: the body mass index (BMI), the degree of airflow obstruction assessed by the forced expiratory volume in one second (FEV1), the modified Medical Research Council (mMRC) dyspnea scale, and the exercise capacity assessed by the 6-min walking distance (6MWD) test.

The secondary outcome measures include polysomnogram, cardiopulmonary exercise test, Beck Depression Scale-II and Saint George’s Respiratory Questionnaire.

1. Polysomnogram (PSG) is performed using the standard PSG (Embletta MPR PG). Electroencephalogram, electrooculograms, chin electromyogram, nasal pressure detected by airflow pressure transducer, respiratory effort, electrocardiography, pulse oximetry and position will be recorded. Sleep indexes include apnea hypopnea index (AHI), longest apnea time (LAT), lowest SaO2 (LSaO2), the percent of the total recorded time spent below 90% oxygen saturation (Ts90%).
2. Cardiopulmonary exercise test (CPET): By evaluating physiological and perceptual responses to CPET in patients, circulatory function, respiratory function can be assessed. The relevant indexes include oxygen uptake (VO_2_%pred), oxygen pulse (VO_2_/HR% pred), oxygen ventilation equivalent (VE/VO_2_ or EQO_2_), carbon dioxide ventilation equivalent (EQCO_2_), respiratory reserve (BR), respiratory exchange rhythm (RER), pulse oximetry (SpO_2_), heart rate (HR) and blood pressure (BP). Oxygen uptake is the main observation. The test was performed with the CS-200 Ergo-Sana Exercise Cardiorespiratory Test Training System from SCHILLER AG, Switzerland.
3. Beck Depression Scale-II: This scale is used to measure depression severity. Patients are required to fill in the scale according to their mood for the last 2 weeks. The score of the test varies between 0 and 63.
4. Saint George’s Respiratory Questionnaire (SGRQ): The SGRQ mainly assesses three conditions including respiratory symptoms (frequency and severity of respiratory symptoms), mobility (effects on and adjustment of everyday activities), and psychosocial impact. A total score is calculated with a maximum of 100 points.

### 2.5 Sample size calculation

This study is a randomized controlled trial. BODE index is the main outcome index, according to the review of literature ^20^, the BODE index of the control group is 4.5±1.3, and it is expected that the BODE index of the treatment group can be decreased by 0.8 by pulmonary rehabilitation, setting the bilateral α=0.05 and the degree of certainty is 90%. The sample size is calculated according to the following sample size calculation formula.

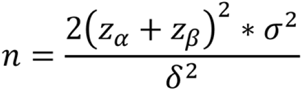

It can be obtained as n=27 cases. Taking into account the 1:1 randomization grouping, 27 cases each of study subjects are required for the intervention and control groups. Besides, considering 20% of missed visits as well as refusals, the final minimum number of study subjects required for the intervention and control groups is 33 cases each, and at least 66 study subjects are included in total.

### 2.6 Randomization and blinding

A statistician will generate a random sequence of 66 numbers using SAS 9.2 software independently, allocating patients to Baduanjin group or the control group in a 1:1 ratio. The random sequence will be put into sealed, opaque envelopes by a doctor not involved with the study to avoid selection bias. Once a patient meets all the criteria for participation, a random number will be delivered by researchers that determines whether the patient will receive Baduanjin exercise or not. Researchers will open an envelope in sequence. Until baseline data have been collected for the first phase of the study, all patients will remain blind. Due to the fact that both the participants and exercise coaches cannot be blinded in this trial, we will blind the outcome assessors and statistics analyzers by the following procedures. Two independent investigators who are not involved in the process of allocation and exercise intervention will be responsible for measuring the outcomes. Prior to the outcome measurement, participants, exercise coaches, and research assistants will be instructed not to disclose any information regarding the allocation. An independent statistician will conduct the statistical analysis, and the allocation information for the treatment groups will be concealed by a blind code. Upon completion of the statistical analysis, the blind code will be revealed.

### 2.7 Data management

The Guangdong Provincial Hospital of Chinese Medicine will supervise the following: randomization of subjects, monitoring of study progress, data management, and statistical analysis. All investigators participated in data management and analysis will be blinded to intervention allocation. All patient data will be collected by trained clinical investigators using standardized paper case report forms (CRFs). CRFs will be locked for revision in preparation for data entry if they are completed. All CRFs are kept in a secure and lock-protected location. To ensure integrity and accuracy of data collection, we perform a double-entry method. In detail, data from the CRF is entered into the database by two investigators independently. Next, a third person (data supervisor) will review the recorded data to ensure consistency and accuracy. If the database records are inconsistent, the data in dispute will be confirmed from the original CRF. If CRF table information is incorrect or missing, the data supervisor will communicate with the clinical investigators and ask them to rectify the error as soon as possible. After verifying the accuracy and integrity, the database will be locked for further analysis.

### 2.8 Safety/security assurance

During the study intervention, participants are monitored weekly for any adverse events defined as any undesirable experience. All adverse events during the study will be recorded on an adverse event CRF and will be evaluated for relevance to the intervention. All adverse events and related treatments will be recorded in the CRF, and the relevance between adverse events and Baduanjin intervention will be evaluated. Adverse events will also be reported to our institutional research ethics boards promptly in accordance with guidelines. Any changes in medical therapy will be documented in CRF, including the date, reasons, and the treatment that followed.

### 2.9 Statistical analysis

Data analysis is conducted using SPSS 18.0 statistical software. For continuous variable, a normality test is performed first. If the results meet normal distribution and the variances within groups are equal, t-test is used between groups; otherwise, the nonparametric Wilcoxon rank-sum test may be used. For categorical variable, chi-squared tests were used for unordered data, and the Wilcoxon rank-sum test was used for ordinal data. Differences are statistically significant at *P*<0.05.

## 3. Discussion

OSA-COPD overlap syndrome was first introduced by David Fenley in 1985, and it described the co-existence of OSAH and COPD. OSA-COPD overlap syndrome is associated with more symptoms and exacerbation events, worse quality of life, and higher mortality compared to either condition alone. Baduanjin exercise combines mind and body to cultivate vital energy, and its movements are gentle, slow, smooth and consistent. Baduanjin is effective in improving balance, leg strength, and mobility and is a safe and sustainable form of home-based exercise for people with chronic stroke^21^. Community based exercise of Baduanjin could help to increase self-efficacy in patients with cardiovascular diseases, thus better self-management of rehabilitation process^22^. Baduanjin exercise provides a safe and feasible treatment option for patients with knee osteoarthritis, as well as offers reductions in pain, stiffness, and disability, which helps improve the patients’ quadriceps strength and aerobic ability^23^. This simple traditional exercise is recommended for Taiwanese patients with heart failure in order to improve their fatigue and quality of life^24^. Baduanjin exercise is an effective and feasible approach to improve self-reported sleep quality but less likely the quality of life in community-dwelling elderly men and women with sleep disturbances^25^.However, its effect for COPD patients with OSAH has not been confirmed by objective evaluation. We conducted this trial to assess whether the addition of Baduanjin to conventional treatment will further improve quality of life.

There are several strengths of our trial. First of all, we studied Baduanjin exercise for the treatment of OSA-COPD overlap syndrome. Secondly, we adopted multidimensional outcome indicators to evaluate pulmonary function and exercise tolerance. In addition, patients can benefit more from the collaboration of medical staff and professional sports coaches. There are still some limitations. As a non-pharmacological treatment, it is difficult to conduct blinding. Therefore, we strictly follow the CONSORT statement requirements to minimize bias. The sample size of this study is still limited, but we will recruit more patients with support in the future. In addition, the patient’s compliance in participating in Baduanjin exercise also deserves our attention. We will establish a good doctor-patient relationship with a professional coach so as to ensure the implementation effect. In summary, this trial will evaluate the effects of Baduanjin exercise in COPD patients with OSAH.

## Trial status

The trial is currently ongoing; we expect patient recruitment to be completed in 2015.

## Data Availability

All relevant data from this study will be made available upon study completion.

## List of abbreviations

IPAP: inspired positive airway pressure
EPAP: expiratory positive airway pressure
CRF: Case Report Form
BODE: B:Body Mass Index;O:Obstruction;D:Dyspnea;E:Exercise Capacity;
SAS: Statistics Analysis System
SPSS: Statistical Product and Service Solutions

## Declaration

### Ethics approval and consent to participate

The study received approval from ethics committee of Guangdong Provincial Hospital of Chinese Medicine (NO. BF2022-139-1). The investigator will explain the benefits and risks of trial participation to patients and their families, and written informed consent should be obtained before patients participate in the trial.

### Consent for publication

Written informed consent was obtained from the patient for publication.

### Availability of data and materials

The data and materials during the current study are available from the corresponding author on reasonable request.

### Authors’ contributions

Xi Zhang and Zhong-De Zhang designed research ; and Yi-hang Cai wrote the paper ; Kang-Jie Ye analyzed data and Xi zhang, Yi-hang Cai and Yu-Jin Li performed research; Ji-Qiang Li contributed new analytic tools.

## Acknowledgements

Thanks to the Guangdong Provincial Hospital of Chinese Medicine, for their guidance in study design and statistics.

